# Cardiometabolic Risk and Diagnostic-Laboratory Reference-Range Disagreement in 794,811 Indian Insurance Applicants: An Aggregate Analysis of Digitized Medical Examinations

**DOI:** 10.64898/2026.06.01.26354654

**Authors:** Saif Lakhani

**Affiliations:** University of California, Davis, CA, USA

**Author notes:** This work was conducted independently and is unrelated to the author’s employment at Amazon.com, Inc. Amazon did not fund, direct, review, or approve this research.

## Abstract

This study analyzes 794,811 digitized medical examinations from Indian life-insurance applicants, a working-age, urban-skewed demographic often undersampled by national surveys. The cohort exhibits a pronounced South-Asian cardiometabolic risk profile: among valid adult records, 41.9% met the criteria for dyslipidemia (driven heavily by low HDL and elevated triglycerides), and 61.4% met AHA 2017 criteria for stage 1 hypertension. However, canonicalizing this dataset across 33,244 diagnostic centers revealed significant heterogeneity in laboratory reference ranges. At the clinical prediabetes threshold of 110 mg/dL for fasting blood sugar, the record-pair disagreement rate across laboratories was 49.7%, with similar variance across other common tests. This structural inconsistency materially affects patient classification and the tracking of disease prevalence, underscoring a critical need for the national standardization of laboratory reporting in India.

## 1 Introduction

The Indian cardiometabolic literature is mature in its general-adult findings. Anjana et al. [1], in the ICMR-INDIAB cross-sectional study across fifteen states, reported adult diabetes prevalence at 7.3% with prediabetes higher still. Gupta et al. [2] summarized adult hypertension at approximately 30% with a sharp urban–rural gradient. Joshi et al. [3], in the ICMR-INDIAB dyslipidemia analysis, documented the South-Asian atherogenic pattern at scale: high triglycerides and low HDL with relatively modest LDL elevation, the same phenotype that McKeigue [4] first described in Bangladeshi adults in east London and that the INTERHEART study [5] ranked as the highest attributable risk factor for myocardial infarction in South Asians.

Less directly measured is the working-age, urban, employed slice of the Indian adult population. NFHS-5 [6] is the standard reference for general-adult prevalence but undersamples the demographic that completes a life-insurance medical examination. The corpus considered here comprises 794,811 examinations accumulated between 2023-01 and 2026-05, processed under data-processing agreements with Indian life insurers.

This paper reports two analyses of that corpus. The first, in Sections 3 and 4, describes cardiometabolic findings: demographics, hypertension, and dyslipidemia. The second, in Section 5, quantifies laboratory-reference-range heterogeneity across the 33,244 diagnostic centers in the dataset and the resulting classification noise at clinical decision values. The second analysis emerged from a canonicalization step required by the first; it is reported in its own right because the magnitude of the disagreement materially affects how the cardiometabolic findings should be read. Section 6 discusses both findings together. Section 7 enumerates limitations in approximate order of importance. The analysis code is open-source at https://tinyurl.com/2cs4w2u7.

## 2 Methods

### 2.1 Data source

The corpus comprises 794,811 medical examinations (797,302 ingested, of which 2,491 were dropped at physiological-plausibility validation; see Section 2.3), conducted between 2023-01 and 2026-05 on behalf of Indian life insurers and processed by an automated extraction pipeline. Each examination originated as a multi-page PDF in one of several thousand layouts. The pipeline runs Mistral OCR (Mistral 2024) for text extraction and Google Gemini 2.5 Pro (Google DeepMind 2024) for structured-data extraction against a fixed JSON schema. Each examination produces a JSON record consisting of a header element, a TESTS element containing categorized lab results, and (when the examiner’s form was completed digitally) a `mer_data` element containing anthropometry, vitals, and a lifestyle questionnaire. Records lacking the `mer_data` element have no anthropometry or lifestyle data by construction; this contributes to the variation in coverage rates reported alongside each analytic result. Across the 794,811 valid records the dataset references 33,244 unique pseudonymized laboratories, 33,182 of which appear in at least one valid record.

The analyses reported here were conducted on the JSON output; source PDFs, OCR intermediate text, and LLM extraction prompts were not accessed during the study.

### 2.2 Anonymization and ethics

All directly identifying fields were excluded at ingestion: patient name, date of birth, identification numbers (Aadhar card, policy, proposal, client), and laboratory contact information. Examination dates were generalized to month and year. Diagnostic-laboratory names were replaced with deterministic pseudonyms (`Lab_0001, Lab_0002, …, Lab_33244`); the original-name-to-pseudonym mapping was retained locally and excluded from all output artifacts, including the manuscript and supplementary materials. No individual-level data appears in any reported result. Cross-tabulations were checked against a *k*-anonymity threshold of 10; sub-groups smaller than 10 were suppressed.

The data were collected under standard insurance underwriting consent.^1^ The data-processing agreements between the processing entity and originating insurer customers permit aggregate, fully de-identified research output as a legitimate use of the processed data. The originating insurer customers were notified prior to manuscript submission with a 10-business-day opt-out window. The manuscript identifies neither the originating insurers nor the commercial customers of the processing pipeline.

### 2.3 Validation, derived fields, and OCR sentinel values

Each record was checked against physiological-plausibility bounds: age 0–120 years, height 50–250 cm, weight 15–300 kg, systolic BP 60–260 mmHg, diastolic BP 30– 160 mmHg. Records failing any check were retained in the parquet output with `is_valid=False` and excluded from analysis. The validation flagged 2,491 records (0.3%) as physiologically impossible. Derived fields included BMI (computed from height and weight, with a sanity gate restricting valid output to 10–80 kg/m^2^), parsed systolic and diastolic blood pressure from the slash-separated input format, sex normalization, and age-bucket assignment.

Two OCR-pipeline conventions required handling. First, the upstream pipeline uses an age value of zero as a missing-value sentinel rather than as a literal age; 50.6% of validated records carried `age=0`. All age-related analyses therefore treat `age=0` as missing, and the adult-age analytic cohort (age ≥ 18 after dropping the sentinels) is 299,607 records.

Second, the upstream pipeline produced over a hundred unique sex strings in development data, including “Mall,” “Mole,” “Mate,” “Femde,” “Femele,” “FeMule,” “MAUS,” and “MVALE,” all clearly OCR-corrupted forms of “Male” or “Female.” A fuzzy-normalization step recovers most of these by mapping short M/N/H-prefixed strings to “Male” and short F-prefixed strings to “Female.” After normalization on the full corpus, the residual unrecognized sex values consist of intentional vocabulary codes (“1,” “2,” “Y/F,” “Y/M,” “Others”) rather than further OCR errors; these were not normalized further.

### 2.4 Examination-date resolution

Examination dates were resolved from three sources in priority order: the JSON header dte field, two filename patterns (a date-prefix pattern used by modern files and a phone-timestamp pattern used by older files), and otherwise null. Resolved dates were assigned to a YYYY-Q[1-4] quarter and a `YYYY-MM` month. Of the 794,811 records, 78% resolved via the header field, 10% via filename, and 12% had no resolvable date.

### 2.5 Statistical methods

Prevalence rates are reported as percentages of the analytic denominator, defined as the subset of records with the relevant field present. Where the denominator differs materially from the full validated cohort, this is reported alongside the rate. Reference-range disagreement, which is not a standard measure, is defined formally in Section 5.1.

Analysis modules are implemented in Python 3.10+ (`pandas, pyarrow, scipy, matplotlib, seaborn`) and verified with 269 unit and property-based tests using pytest and hypothesis. The analysis is reproducible from the parquet files and the public analysis-module repository.

### 2.6 Scope

The study makes no individual-level claims. It does not link any examination to any individual, laboratory, or insurer. It does not address insurance underwriting decisions, claims outcomes, or policy pricing. The population is “working-age Indian life-insurance applicants who completed a digitized medical examination between 2023-01 and 2026-05” and has known biases (urban-skewed, employed-skewed, predominantly male in this cohort) which are detailed in Section 7.

### 2.7 Use of Artificial Intelligence

During the preparation of this manuscript, Google Gemini 2.5 Pro (via the Gemini CLI tool) and Anthropic Claude Sonnet 4.6 (via the Claude Code CLI tool) were used to assist in revising the prose, refining the narrative tone of the text, and developing analysis code. The AI tools were not used in the conceptualization of the research, the generation of data, or the statistical analysis. The author manually reviewed, fact-checked, and edited all AI-assisted output, and takes full responsibility for the final content and integrity of the publication.

## 3 Demographics

The median age in the adult-age cohort (*n* = 299,607) was 38 years (IQR 30–47). The cohort was 68.6% male and 31.4% female, with the remainder in non-resolvable or non-binary categories that were excluded from sex-stratified analyses. After accounting for the upstream OCR sentinel value (`age=0` treated as missing per Section 2.3), age coverage was 37.7%; sex coverage was 93.4%. The male skew is consistent with the demographics of Indian life-insurance applications, in which men are the dominant policy buyers; NFHS-5 [6] reports a similar skew in the insurance-purchasing subpopulation.

Anthropometry was available on a smaller analytic cohort. Of records with both height and weight extracted (*n* = 103,597, BMI coverage 13.0%), the mean BMI was 26.1 kg/m^2^ and the median was 25.7 kg/m^2^. Under WHO categorization, 41.9% of these records were overweight (BMI 25.0–29.9) and 15.6% were obese (BMI≥ 30); the combined fraction above the WHO normal range exceeded half of the BMI-having subset.

The 13.0% BMI coverage warrants explicit attention. Heights and weights were inconsistently extracted by the upstream OCR pipeline, particularly when handwritten on the medical examiner’s form, and records following the 2-element JSON variant did not contain a `mer_data` block and therefore no anthropometry by construction. The BMI-having subset of 103,597 records is therefore both substantial in absolute terms and a non-random sample of the cohort; the bias risk is addressed in Section 7.

The Asian-Pacific consensus on BMI categorization [9] argues that South Asians develop cardiometabolic complications at lower BMI than European populations and recommends an obesity threshold of 27.5 kg/m^2^ rather than 30. Application of the lower threshold yields a substantially higher obese fraction in the present cohort. The choice between WHO and Asian-Pacific cutoffs is a guideline-setting question outside the scope of this paper; the data are consistent with the observation that the WHO cutoff may under-detect cardiometabolic risk in South Asian populations [9].

## 4 Cardiovascular findings

### 4.2 Hypertension

The medical-examiner’s form records up to three blood-pressure measurements (BP1, BP2, BP3); analyses use BP1, the first recorded reading, due to inconsistent extraction of subsequent readings. Of the 794,811 records, 291,552 (36.7%) had a parseable BP1 value.

Under AHA 2017 categorization [7], 27.3% of BP-having records were normal (systolic < 120 and diastolic < 80), 11.3% were elevated (systolic 120–129 and diastolic < 80), 43.4% were stage 1 hypertension (systolic 130–139 or diastolic 80–89), 17.3% were stage 2 hypertension (systolic ≥ 140 or diastolic ≥ 90), and 0.7% were in hypertensive crisis (systolic ≥ 180 or diastolic ≥ 120). Combined stage 1 or higher prevalence was 61.4%.

The reported AHA-2017 prevalence requires contextualization. Under the older JNC 7 cutoffs (systolic ≥ 140 or diastolic ≥ 90), the prevalence in the same cohort is 18.0%, comparable to the 21% adult prevalence reported by NFHS-5 [6] and the approximately 30% summary figure in Gupta et al. [2]. The AHA 2017 reclassification, which India has not formally adopted, increases apparent hypertension prevalence approximately threefold relative to JNC 7. The choice of cutoff is therefore consequential rather than nominal: the gap between 18.0% and 61.4% represents a reclassification decision that, if adopted at policy level in India, would affect tens of millions of working-age adults.

The age and sex gradients of hypertension prevalence are monotonic and consistent with prior literature. Among males, AHA stage 1+ prevalence rose from 49.2% in the 18–24 age band to 78.1% in the 65+ band. Among females, the corresponding range was 31.4% to 77.4%. The male– female gap was largest in young adulthood and narrowed substantially with age.

### 4.2 Dyslipidemia

Lipid coverage substantially exceeded BP coverage: 99.5% of validated records had at least one lipid value, compared to 36.7% with BP. The difference reflects procedural conventions of insurance medical examinations (lipid panels are nearly always ordered) and the higher reliability of OCR extraction from typed laboratory printouts than from handwritten examiner forms.

Test-name surface forms were canonicalized into four primary markers (total cholesterol, LDL, HDL, triglycerides). VLDL was excluded as it is not in the ATP III classification scheme [8], and lipid ratios were excluded as derived rather than measured quantities. Per ATP III thresholds:

- Total cholesterol ≥ 200 mg/dL: 15.7% of the 789,574 records with a measurement.
- LDL cholesterol ≥ 130 mg/dL: 10.3% of the 430,123 records with a measurement.
- Sex-specific low HDL (< 40 mg/dL male, < 50 mg/dL female): 35.2% of the 497,573 records with both HDL and a known sex.
- Triglycerides ≥ 150 mg/dL: 20.6% of the 552,665 records with a measurement.
- At least one of the above (any-dyslipidemia): 41.9%.

The pattern observed is consistent with the South-Asian atherogenic-dyslipidemia phenotype reported by McKeigue [4] and Joshi et al. [3]. Low HDL was the highest-prevalence single marker at 35.2%, followed by elevated triglycerides at 20.6%; total cholesterol and LDL elevations were comparatively modest at 15.7% and 10.3%. The 41.9% any-dyslipidemia rate falls within the 30–40% range reported by Joshi et al. [3] for working-age Indian cohorts. INTERHEART [5] identified this lipid pattern as the highest-attributable-risk metabolic profile for myocardial infarction in South Asians.

Figure 3 shows the four lipid distributions with ATP III threshold lines indicated.

**Figure 1:**
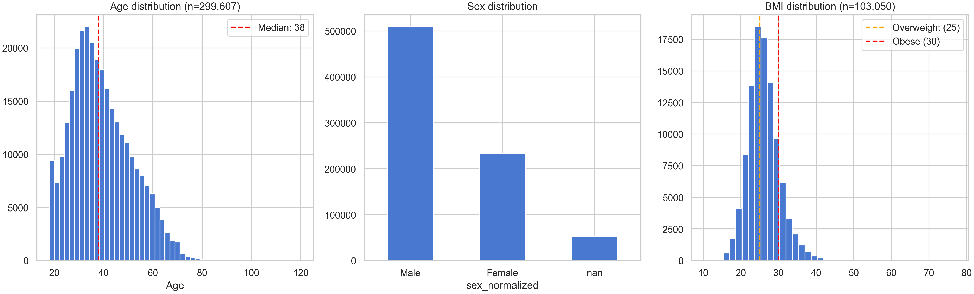
Age, sex, and BMI distributions for the validated cohort.

**Figure 2:**
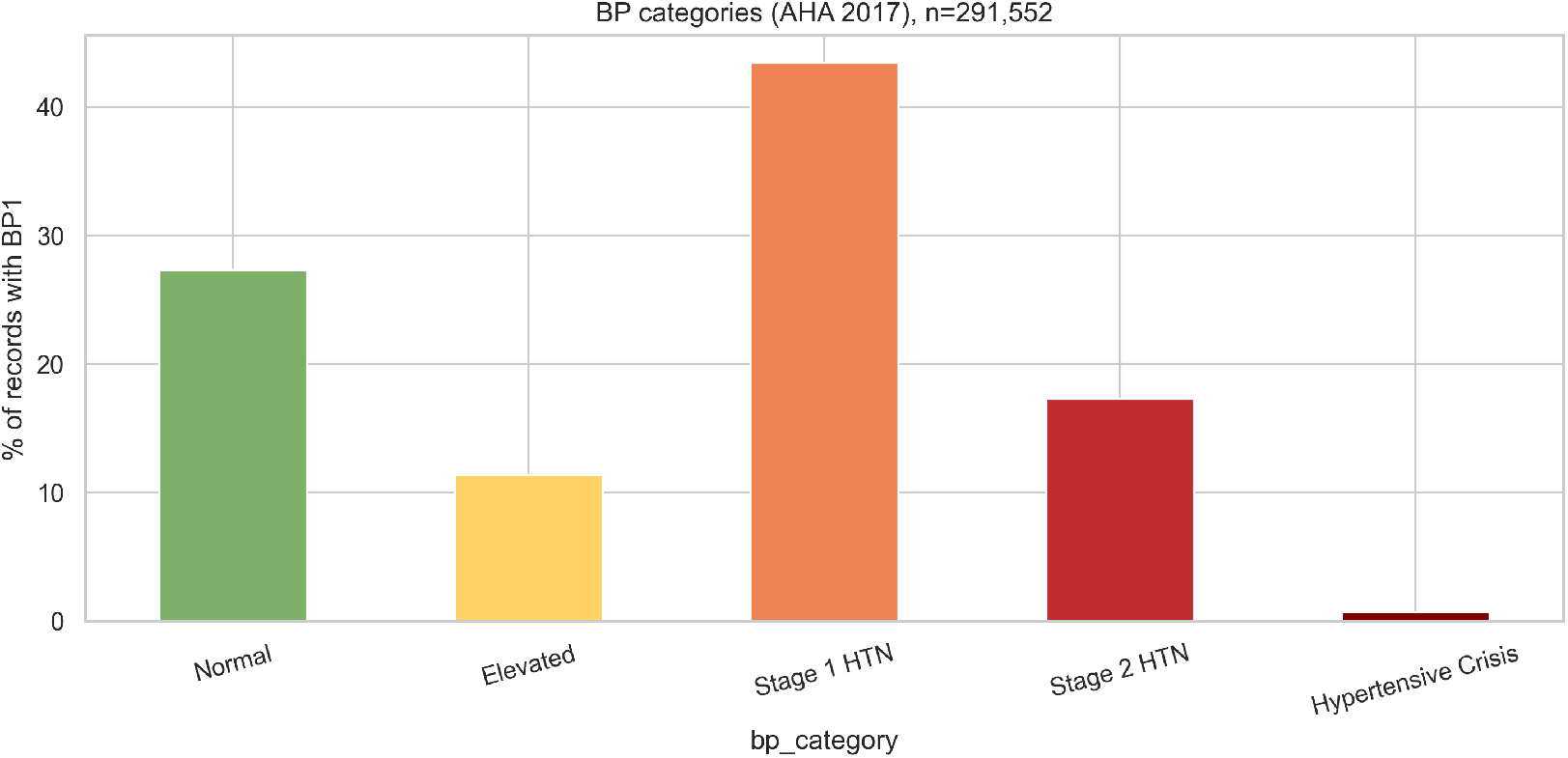
Blood-pressure category breakdown by AHA 2017 classification (left) and prevalence of stage 1+ hypertension by age band and sex (right).

**Figure 3:**
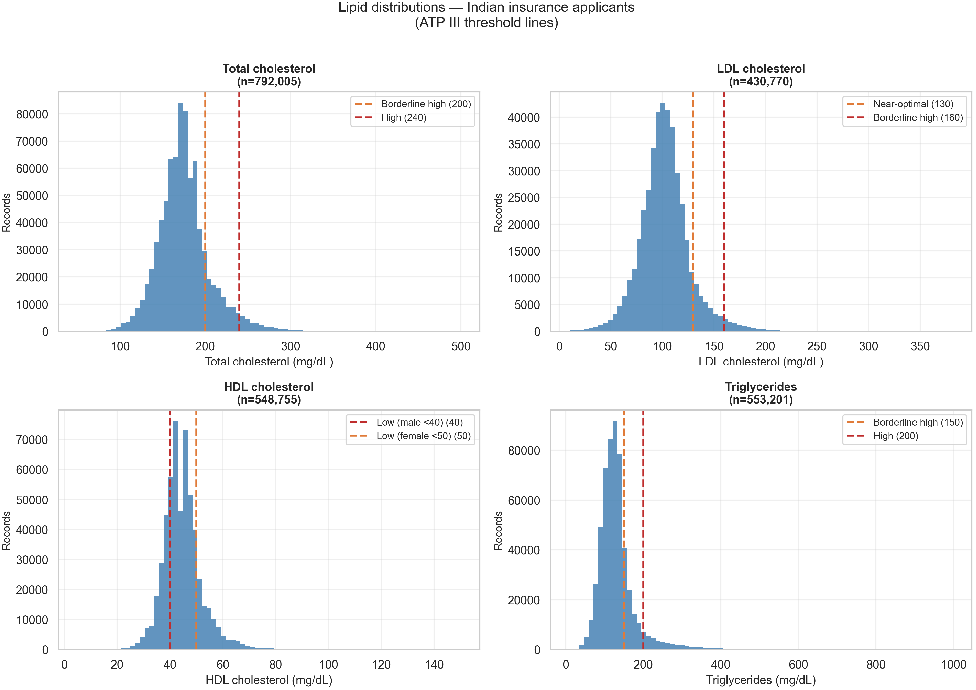
Distributions of total cholesterol, LDL, HDL, and triglycerides with ATP III threshold lines.

## 5 Laboratory-reference-range disagreement

This analysis was not part of the original study aims. A canonicalization step required for the cardiometabolic analyses revealed substantial heterogeneity of reference ranges across the 33,244 diagnostic laboratories in the corpus. The magnitude of the heterogeneity, and its implications for the cardiometabolic findings reported in Sections 3 and 4, justify reporting it as a primary analytic result.

### 5.1 Definition

Each laboratory in the corpus prints its own reference range for every test. Diagnostic interpretation at the report level depends on the reference range the laboratory printed: a serum SGPT of 45 U/L is classified as elevated under a reference upper limit of 40 U/L and as normal under an upper limit of 50 U/L, the same numeric value receiving different classifications depending on the producing laboratory.

The pairwise reference-range disagreement rate at a given clinical decision value is defined as the fraction of record pairs whose reference upper limits classify that value differently. Formally, for *N* records on test *T* with reference upper limits {*mi*}, and a clinical decision value *v*, the disagreement rate is the count of unordered pairs (*i, j*) for which *mi* < *v* ≤ *m j* (or vice versa) divided by 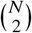. The “weighted by record volume” version, reported here as the primary measure, treats every record as one unit of the pair denominator, making the resulting rate population-level rather than lab-pair-level.

Three preprocessing steps were required. First, test-name surface forms were canonicalized (e.g., “SGPT,” “ALT,” “ALT/SGPT,” “Alanine Aminotransferase,” “Alanine aminotransferase (ALT)” all map to canonical key `sgpt`). Second, reference-range OCR artifacts were filtered using per-test plausibility bounds (negative upper limits, values exceeding plausible bounds, swapped min/max). Third, clinical decision values were selected from ATP III and KDIGO guidelines for the lipid and kidney measures, WHO/ICMR criteria for fasting blood glucose (110 mg/dL impaired fasting glucose threshold), and conventional clinical cutoffs for SGPT, SGOT, GGT, and hemoglobin.

### 5.2 Results

For ten common tests at their clinical decision values, weighted by record volume, see Table 1. Mean disagreement across the ten tests: 24.8%. Maximum: 49.9%, on LDL at 130 mg/dL (the ATP III borderline-high threshold); fasting blood sugar at 110 mg/dL was 49.7%; SGOT at 40 U/L and GGT at 50 U/L each reached 46.3%.

**Table 1:** Reference-range disagreement at clinical decision values. Population-weighted record-pair disagreement across the ten high-volume canonicalized tests.

| Test | Clinical value | *n* records | Distinct upper limits | Upper-limit IQR | Disagreement % |
| --- | --- | --- | --- | --- | --- |
| LDL | 130 mg/dL | 296,916 | 68 | 100–145 | **49.9** |
| Fasting blood sugar | 110 mg/dL | 363,059 | 46 | 100–110 | **49.7** |
| SGOT (AST) | 40 U/L | 503,024 | 86 | 37–41 | **46.3** |
| GGT | 50 U/L | 400,278 | 109 | 40–55 | **46.3** |
| Creatinine | 1.2 mg/dL | 249,378 | 53 | 1.2–1.4 | **28.0** |
| SGPT (ALT) | 40 U/L | 568,005 | 93 | 40–45 | **23.1** |
| Triglycerides | 150 mg/dL | 473,421 | 62 | 150–165 | 3.3 |
| Total cholesterol | 200 mg/dL | 559,502 | 54 | 200–200 | 1.7 |
| HDL | 45 mg/dL | 482,702 | 63 | 60–70 | 0.1 |
| Hemoglobin | 12 g/dL | 409,829 | 69 | 16–17 | 0.0 |

Figure 4 shows the disagreement curve as a function of patient value for each test. LDL shows a broad plateau of near-maximum disagreement across its full IQR (100–145 mg/dL), reflecting genuine dispersion of Indian-laboratory upper limits across that range. GGT, SGOT, and FBS show narrower peaks centered on their clinical decision values. Total cholesterol and triglycerides are outliers in the opposite direction: the standard ATP III thresholds of 200 and 150 mg/dL have effectively converged across Indian laboratories, producing near-zero pairwise disagreement at those values.

**Figure 4:**
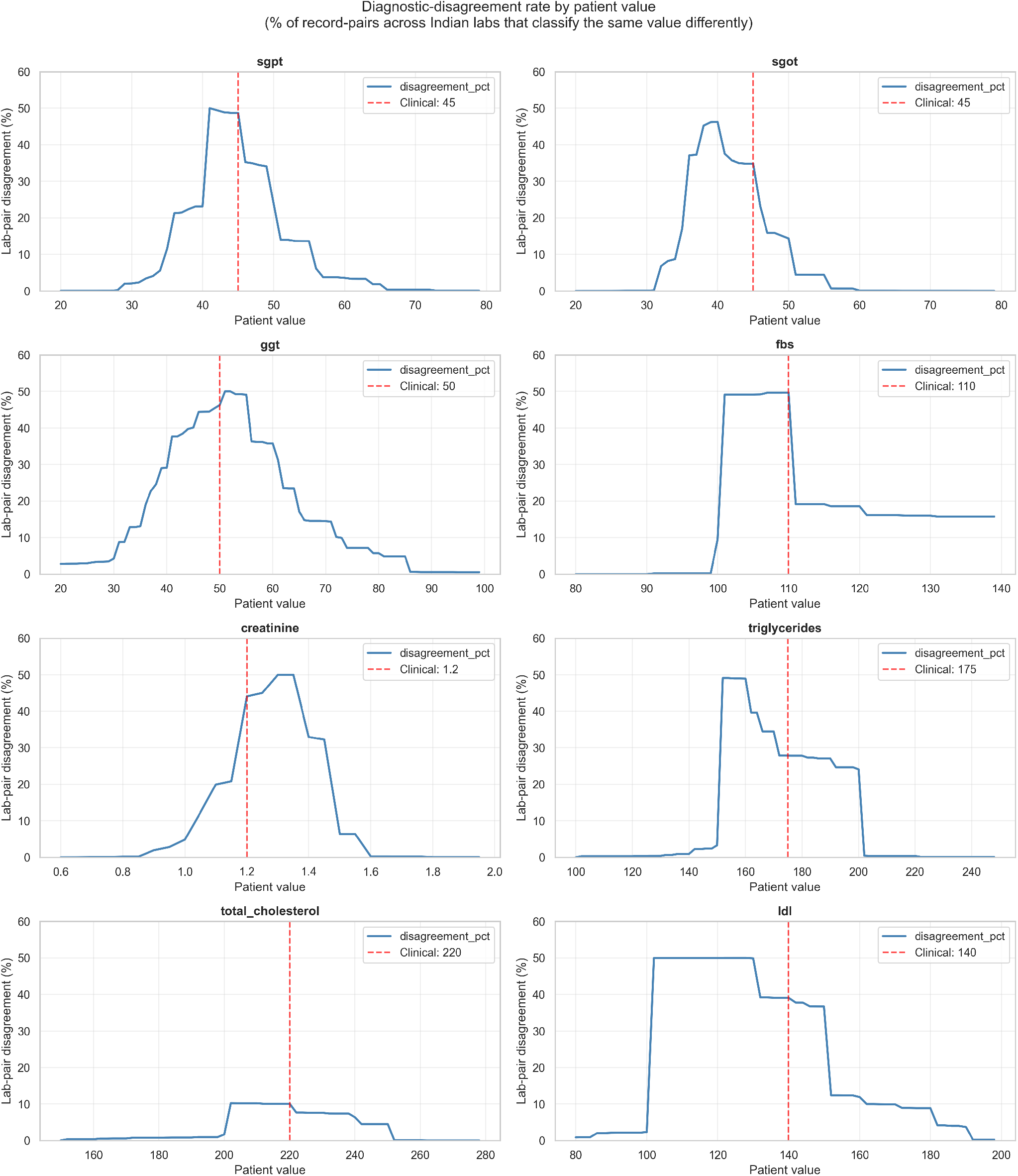
Laboratory-reference-range disagreement curves for high-volume tests at clinical decision values.

The disagreement reflects classification heterogeneity rather than assay heterogeneity. Indian diagnostic laboratories measure SGPT to comparable analytic precision; the interpretation of the same numeric value via the reference range printed on the report is what differs.

### 5.3 OCR artifacts and within-laboratory variation

Reference-range OCR artifacts in the raw data included negative upper limits, upper limits in the thousands (probable comma misplacements), and zero values used as missing-value sentinels. The disagreement rates reported in this section are computed after filtering these using per-test plausibility bounds. The 49.9% disagreement on LDL, 49.7% on FBS, and 46.3% on SGOT and GGT are not residual OCR error; they are heterogeneity in the published reference ranges.

Within-laboratory reference-range variation across records (which would contaminate the inter-laboratory analysis) was uncommon. Of the 33,244 unique laboratories, the median laboratory used a single reference upper limit per test in the ten-test set examined.

### 5.4 Test-name and unit-string heterogeneity

Across 20,787,138 lab-result rows, the corpus contained 9,165 distinct `test_name` strings and 21,223 distinct unit-string representations. For a domain comprising at most a few hundred genuinely distinct tests and approximately fifteen distinct units, this represents a vocabulary-control problem analogous to those addressed internationally by LOINC for test naming and UCUM for units. No equivalent Indian standard currently exists. The canonical map released as Supplementary Material 1 covers the ten high-volume tests reported in Section 5 and accounts for 24.0% of the 20.8M lab-result rows; the long tail of less-common test names is not yet canonicalized.

## 6 Discussion

### 6.1 The cardiometabolic findings

The cardiometabolic results reported in Sections 3 and 4 are consistent with the recent Indian literature, on a single-source cohort that exceeds prior comparable studies in size. Median age 38 years, 68.6% male, mean BMI 26.1 kg/m^2^, any-dyslipidemia 41.9%, low HDL 35.2%, elevated triglycerides 20.6%, AHA stage 1+ hypertension 61.4% (or JNC 7 stage 1+ at 18.0%): these findings confirm an established pattern with smaller statistical error bars than prior single-source studies in this demographic afforded.

The South-Asian atherogenic-dyslipidemia phenotype, first described by McKeigue [4] and confirmed at scale in INTERHEART [5], is visible in the data. Low HDL is the most prevalent single dyslipidemia marker at 35.2%, elevated triglycerides at 20.6%, with elevated total cholesterol and LDL substantially less common. This is the expected ordering for the South-Asian phenotype, not the LDL-dominant ordering observed in European cohorts.

### 6.2 The reference-range-disagreement findings

The reference-range-disagreement results reported in Section 5 carry direct implications for the interpretation of the cardiometabolic findings. The any-dyslipidemia prevalence of 41.9% is meaningful insofar as “dyslipidemia” represents a consistent classification across the laboratories that produced the underlying numeric values. For total cholesterol at 200 mg/dL and triglycerides at 150 mg/dL, where Indian-laboratory reference ranges have effectively converged on the ATP III thresholds, the classification consistency is high. For LDL at 130 mg/dL, where the disagreement rate is 49.9%, two random Indian-laboratory records of the same numeric LDL value would receive different classifications in approximately one of every two pairs. Reported prevalence rates for laboratory-flagged abnormalities therefore carry classification noise that varies systematically by test.

Three implications follow.

#### Clinical practice

Flagged abnormal results on tests with high inter-laboratory disagreement (LDL, FBS, SGOT, GGT, and SGPT) should ideally be re-evaluated against the patient’s own historical laboratory’s reference range when feasible. Where this is not feasible, clinicians and primary-care decision-support systems should treat single-laboratory abnormality flags on these tests as preliminary rather than definitive.

#### Insurance underwriting

Risk classifications derived from these laboratory values carry classification noise that is invisible to the underwriter. An applicant routed to one laboratory versus another may receive different classifications on identical biology. Actuarial models that incorporate laboratory-flagged risk markers should consider modeling the inter-laboratory reference-range distribution as an additional noise source.

#### Public-health surveillance

Prevalence rates that aggregate laboratory-flagged abnormalities across heterogeneous Indian laboratories carry the disagreement as additive bias whose direction cannot be determined without harmonization. The Indian Council of Medical Research’s National Reference Range project is the relevant policy lever; the data here supply empirical motivation for its acceleration.

### 6.3 The methodological contribution

The canonicalization mapping constructed for these analyses covers the ten high-volume tests reported in Section 5, which together account for 24.0% of all lab-result rows in the corpus. The remaining surface-form long tail is not yet canonicalized; for the canonicalized subset, the compression from surface form to canonical key is roughly two orders of magnitude. Production systems consuming Indian laboratory data must perform such canonicalization. Until now this work has been undertaken privately and inconsistently across organizations. The canonical map and the laboratory-pseudonymization index from this study are released as Supplementary Materials 1 and 2 to support reuse.

## 7 Limitations

The following limitations are listed in approximate order of importance to the interpretation of the results.

### Coverage variability across analytic fields

The analytic denominator differs by analysis: 794,811 for demographics, 299,607 for adult-age statistics, 291,552 for hypertension (BP coverage 36.7%), 790,961 for at-least-one-lipid, 103,597 for BMI (13.0%), and 497,573 for sex-stratified low HDL. The analytic *n* is reported alongside every result; readers should track which *n* applies to which finding rather than treating all reported rates as derived from the full validated cohort.

### Selection bias of the insurance-applicant population

This cohort is not a random sample of Indian adults. It is urban-skewed, employed-skewed, and male-skewed (68.6%). Cardiometabolic findings describe this specific subpopulation rather than Indian adults at the general population level.

### OCR-pipeline artifacts

The corpus is the LLM-extracted output of an OCR pipeline. Two artifact classes were documented and corrected: `age=0` as a missing-value sentinel, and OCR-corrupted sex strings. Other artifacts likely persist. Source PDFs were not available for validation; the JSON corpus is treated as the empirical data.

### Cross-sectional design

A single examination per applicant, with no follow-up, no claims data, and no clinical outcome data. Cross-sectional findings cannot speak to causation, progression, or prognosis.

### Insufficient Variant A coverage for internal validation

A small fraction of the corpus (0.7%) carries QC metadata from an older variant of the upstream pipeline, in principle usable as an internal validation set. At 0.7% coverage, the Variant A subset is too small to anchor any cross-validation claim; this study did not pursue that direction.

### Reference-range analysis depends on upstream OCR extraction

The reference-range-disagreement analysis relies on the upstream pipeline’s parsed `applicablemin` and `applicablemax` values. Where these are misextracted, the disagreement rate is biased; the bias direction cannot be quantified without re-OCR of the source PDFs. Per-test plausibility filters drop the obvious extraction errors; subtler errors persist.

### No within-laboratory temporal analysis

Whether individual laboratories changed their reference ranges over the 2023-01 to 2026-05 window is not addressed in this study. If laboratories migrated to ICMR-recommended ranges over time, the pooled disagreement rates conflate inter-laboratory and intra-laboratory temporal heterogeneity.

## 8 Conclusions

Working-age Indian insurance applicants in this cohort carry the cardiometabolic risk profile that the recent Indian literature has predicted, on a single-source sample that exceeds prior comparable studies in size. The South-Asian atherogenic-dyslipidemia phenotype is visible. Hypertension prevalence is high under contemporary AHA cutoffs and at the prior literature’s expected level under the older JNC 7 cutoffs, with the gap between the two reflecting a substantive reclassification decision that India has not formally taken. The same corpus reveals reference-range heterogeneity across Indian diagnostic laboratories of a magnitude that materially affects the classification of patients at clinical decision values for several common tests. The findings argue for both clinical attention to the cardiometabolic burden in this population and for accelerated national standardization of laboratory reference reporting.

## Data Availability

All data produced in the present study are available upon reasonable request to the authors. Individual-level data cannot be released; the corpus is held under data-processing agreements that do not permit individual-level publication. Aggregate statistics and the canonicalization map are released alongside the preprint at https://tinyurl.com/2cs4w2u7.

## 9 Data and code availability

The analysis code, including the ingestion pipeline, the canonicalization map, the analysis modules, and the 269-test test suite, is available at https://tinyurl.com/2cs4w2u7. The aggregate-statistics file paper/stats.json and the supplementary canonicalization map are released alongside the preprint. Individual-level data are not, and will not be, released; the corpus is held under data-processing agreements that do not permit individual-level publication. Researchers interested in replicating analyses on a comparable corpus may contact the corresponding author about establishing a similar data-processing agreement.

## Ethics statement

The data were collected under standard insurance underwriting consent. The data-processing agreements between the processing entity and originating insurer customers permit aggregate, fully de-identified research output as a legitimate use of the processed data. No individual-level data appears in any reported result. No ethics committee approval was sought as the study uses only aggregate, fully anonymized statistics derived from routinely collected commercial data with no patient contact, intervention, or identifiable health information in any output.

## Footnotes

1 Formal Institutional Review Board (IRB) review was not required. The study relies exclusively on retrospective, aggregate, and fully anonymized statistics derived from commercial data, with no patient contact, intervention, or access to identifiable health information, meeting standard criteria for exemption from human subjects research oversight.

